# Targeted neural stimulation congruent with immersive reality decreases neuropathic pain – a Randomized Controlled Trial

**DOI:** 10.1101/2024.12.10.24318374

**Authors:** Giuseppe Valerio Aurucci, Noemi Gozzi, Markus Wagner, Greta Preatoni, Nicola Brunello, Natalija Secerovic, Carl Moritz Zipser, Stanisa Raspopovic

**Affiliations:** Neuroengineering Laboratory, Department of Health Sciences and Technology, ETH Zürich; Tannenstrasse 1, 8092 Zürich; Department of Neurology and Neurophysiology and Spinal Cord Injury Center, Balgrist University Hospital, University of Zurich, Zurich, Switzerland; Center for Medical Physics and Biomedical Engineering, Medical University of Vienna, 1090 Vienna, Austria; Institute Mihajlo Pupin, Volgina 15, Belgrade, Serbia

**Keywords:** pain, neurostimulation, VR, biomarkers, EEG, neuropathy

## Abstract

**Background:** Chronic neuropathic pain is a complex experience, posing a major challenge in personalizing its treatment. Present treatments consist of non-specific, standardized drugs that are often addictive, leaving many patients non-respondent and with significant side effects. Designing individualized therapies requires targeting the multidimensionality of pain and developing objective endpoints to demonstrate their effectiveness. Currently, non-pharmacological alternatives are emerging, such as neurostimulation and Virtual Reality (VR), activating pain relief via peripheral neuromodulation and attention modulation. Similarly to drugs, many neurostimulation approaches are unspecific, targeting areas near the pain site and disregarding the neural pathway of pain. Above all, neurostimulation and VR are yet to be evaluated as a combined synergistic intervention, particularly in a randomized controlled trial (RCT).

**Methods and Findings:** To this aim, we developed a targeted neurostimulation congruent with immersive VR platform providing a multisensory pain intervention through the synergistic application of somatotopic electro-tactile and visual stimuli. The endpoints included measuring sensory, neurophysiological (EEG), and self-reported indicators of pain. We tested the efficacy of the multisensory intervention against the control consisting of VR-only intervention on four consecutive intervention days in an RCT (N=18 neuropathic patients). The multisensory intervention resulted in a clinically significant reduction of pain (>50%), lasting up to one-week follow-up. The provided analgesic effect was statistically stronger compared to the VR-only control across treatment days and at follow-up. The clinically relevant pain decrease was accompanied with objective improvements in tactile acuity, proprioceptive measures, and changes in EEG pain biomarkers for the multisensory intervention group only.

**Conclusions:** The developed multisensory treatment showed a clinically significant reductions in self-reported pain, supported by improvements in objective sensory and neurophysiological measures. These results represent a significant advancement in the treatment and assessment of pain, offering a non-invasive, accessible, and cost-effective solution for neuropathic pain, a major societal burden and one of the most prevalent neurological conditions worldwide.

**Clinical trial registration:** The trial was registered with ClinicalTrial.gov (NCT05483816).

## INTRODUCTION

Chronic neuropathic pain presents a major health and economic burden for the society, affecting between 6.9% and 10% of the population ^40,70–72,1,2^. It is caused by a lesion or dysfunction of the peripheral or central nervous system ^3^ and originates from prevalent medical conditions such as diabetes, cancer, infections, spinal cord injury, and stroke ^1,4^. Independently from the etiology, the symptoms are shared across clinical populations, including sensory nociceptive components (burning, shooting, and uncomfortable sensations^5^) and subjective factors. Indeed, alongside the sensory bottom-up components, chronic pain accounts for top-down emotional and cognitive factors directly modulating the descending pain pathways ^6,7^. This complex and multifaceted interplay of different pain dimensions poses a significant challenge to developing effective therapies. As of today, non-specific pharmacological intervention remains the most widely adopted approach ^8^. Yet, antidepressants and anticonvulsants (as a first-line treatment ^9^), and analgesic opioids (as a second-line treatment ^9^) often lead to strong side effects ^10^, addiction and overdose problems ^11^. The social impact of opioid overuse is indeed critical: the rate of overdose deaths linked to prescribed opioids in the US quadrupled between 1999 and 2016 and continues to rise ^12^. Moreover, the efficacy of opioids is often only partial ^13^, as evidenced by the persistently high prevalence of pain over time ^14^, limited long-term analgesic effects ^14–16^, and unsatisfaction with treatment ^17^.

To this end, non-pharmacological alternatives have emerged, including peripheral non-invasive neurostimulation such as Transcutaneous Electrical Nerve Stimulation (TENS). Neurostimulation induces peripheral analgesia by multifactorial processes^18^, triggering the pain-gate mechanism proposed by Melzack and Wall^19^: the stimulation of large-diameter sensory fibers elicits inhibitory mechanisms, blocking the processing of nociceptive information. Despite its clinical potential, at present, the efficacy of neurostimulation is inconclusive due to the heterogeneity of studies, the lack of randomized controlled trials, and the low fidelity of diverse stimulation techniques ^18,20^. Among others, the variability of multiple stimulation parameters (i.e. electrode location, pulse intensity, and pulse width) adopted in clinical studies ^20^ prevents to state its consistency against neuropathic pain ^18^. Moreover, most studies omit a crucial calibration step to determine optimal stimulation parameters for each subject, instead using non-personalized settings regardless of the patient’s neuropathy status (e.g. sensory loss, sensibility threshold). Electrodes positioning, in particular, is critical ^20^ to efficiently activate the fibers of interest ^21^. Most clinical studies ^18^, or commercial devices ^22,23^, use unspecific stimulation sites (often described as ‘near the pain site’), disregarding the anatomical location of the nerves and therefore possibly undermining the full potential of neurostimulation of these fibers.

Alongside neuromodulation, VR has been employed to provide neuropathic pain analgesia by modulating patients’ attention, with the majority of applications focusing on spinal cord injury cohorts ^24,25^. VR has proven effective in altering the subject’s sense of ‘self’ ^26^, broadly defined as body representation^27^. Given the neural connection between body representation and the processing of noxious stimuli ^28^, VR has been advocated as a tool to modulate body perception and alleviate pain^29,30^, but supportive clinical evidence is missing. Moreover, both VR and neurostimulation present the shortcoming of targeting a single aspect of pain (either sensory for neurostimulation or attentional for VR) without addressing its multifaceted nature. In our previous work, we tested the combined use of VR and neurostimulation in a one-day intervention for neuropathic patients ^31^. However, the focus of the previous was on real-time pain recognition rather than on robust therapeutic insights, which were further prevented by the absence of a control condition and the short duration of the intervention. Furthermore, the inherent limitations of current interventions are further amplified by the absence of objective and reliable indicators of therapeutic effectiveness^32,33^. The gold standard for pain assessment relies on self-reports and unidimensional scales, which are inherently subjective and fail to capture the complexity of the pain experience ^34^. Hence, healthcare providers and national agencies advocate for the search for complementary physiological and electrophysiological pain biomarkers to monitor therapeutic response and disease progression ^32,35^.

The limitations of current therapeutic options emphasize the necessity of investigating targeted non-pharmacological interventions and developing methods to thoroughly validate their effectiveness in reducing neuropathic pain. To these aims, we developed a multisensory pain intervention encompassing immersive VR and targeted neurostimulation (tSTIM) and tested its efficacy in a randomized clinical trial (RCT) with eighteen chronic neuropathic pain participants. The intervention spanned four consecutive days of therapy, with participants randomly assigned to either the VR+ tSTIM group (receiving synchronous visuo-tactile stimulation) or the VR group (active comparator, receiving VR stimulation only). To fully validate the impact of the intervention, its efficacy was multimodally assessed by integrating subjective measures of the Neuropathic Symptom Scale Inventory (NPSI)^36^ and Visual Analogue Scale (VAS)^37^, with objective physiological and electrophysiological endpoints. To this end, monitoring of the therapeutic response included measurements of tactile acuity (Two-Point Discrimination Test ^38^), body representation (Proprioceptive Displacement Test ^39^), and electroencephalography (EEG) neuro-correlates of pain ^40^. This controlled, multimodal approach allowed us to objectively validate the intervention as a safe, non-invasive, and effective solution, addressing the shortcomings of current therapeutic methods.

## METHODS

### Participants recruitment

Participants were recruited through the Department of Neurology and Neurophysiology at Balgrist University Hospital, University of Zurich, Switzerland. Inclusion and exclusion criteria are specified in Table S1. 25 participants were preliminary assessed for eligibility. Participants who did not meet all the inclusion criteria were discarded (See CONSORT diagram, Fig. S1). A total of 18 participants participated in the study. All participants read and signed the informed consent form including the use of identifiable images and access to medical records The experiments were designed and conducted in accordance with the Declaration of Helsinki and received approval by the Kantonale Ethikkommission Zürich (Nr. 2021–02258). The trial was registered with ClinicalTrial.gov (NCT05483816).

### Multisensory platform

We developed a multisensory platform (Fig. 1) to deliver non-invasive targeted neurostimulation real-time matched with the visual VR stimulus. The technological framework combines a VR headset (HTC VIVE Pro) with a neurostimulation device (RehaMove3 Hasomed Gmbh) (Fig. 1A). The platform was designed similar to previous studies ^31,78,79^. To collect neurophysiological signature of pain, a 24-channel portable EEG device (SMARTING MOBI, mBrain Train) with a sampling frequency of 500 Hz was employed (Fig. 1A). The VR environment was developed in Unity 3D (version 2019.4) and consisted of a beach scenario where participants saw themselves from a first-person perspective (both male and female avatars were available to further enhance immersion) while sitting on a beach chair in front of the sea (Fig. 1B). During the intervention, participants’ virtual feet were repeatedly touched by incoming waves (Fig. 1B). Synchronously, targeted neurostimulation was provided at the tibial and peroneal nerves of both feet to elicit a touch-like sensation spreading over the plantar and dorsal side of the feet respectively (Fig. 1B). To foster the immersive experience, the pulse width of the electrical stimulation was modulated following a Gaussian curve reaching its maximum at the maximum wave height (Fig. 1B, Fig 1C).

**Figure 1.**
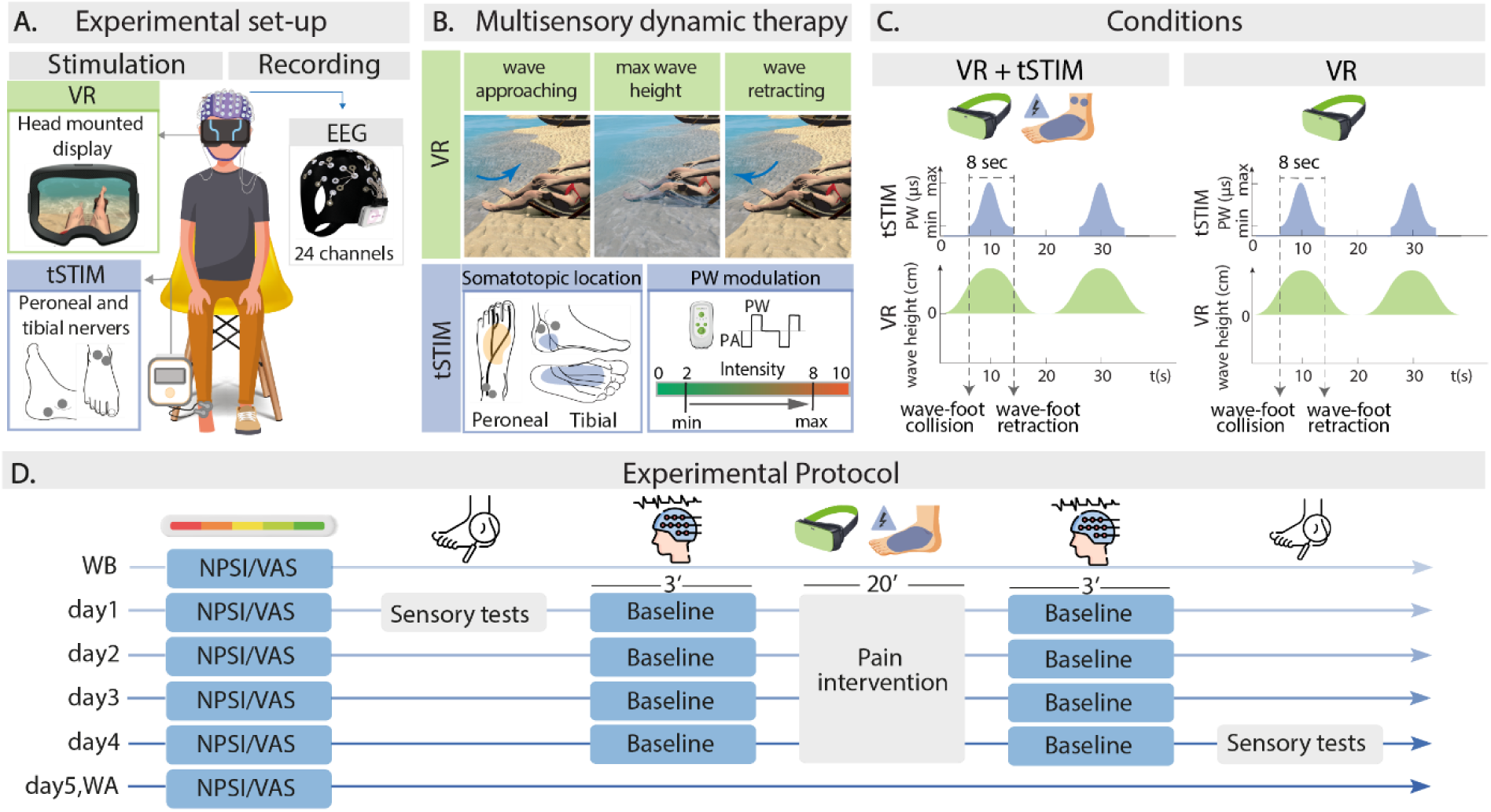
Experimental set-up and protocol of multisensory pain intervention. A) Experimental set-up. Subjects wore a VR headset and received neurostimulation at the peroneal and tibial nerves. For baseline recordings, a 24-channel EEG system was used. B) Pain intervention. The VR stimulation consisted of virtual waves touching the avatar’s feet (wave approaching, reaching its max height, and retracting). tSTIM was delivered at peroneal and tibial nerves to elicit a synchronous sensation spreading over the dorsal and plantar side of the foot respectively. Its pulse width was modulated from a low (2/10) to strong (8/10) sensation to match the height of the wave (Video S1). C) Conditions. The VR+tSTIM group repeatedly received multisensory synchronous stimulations. tSTIM started at the wave-foot collision, followed a Gaussian curve, and ended at the wave-foot retraction (for a total of 8 seconds). The VR group received VR stimulation only. D) Experimental protocol. For four consecutive days, participants underwent 20 minutes of pain intervention. Before and after each intervention, Baseline EEG was collected. On the first and last day of intervention sensory tests were performed. NPSI and VAS questionnaires were collected at the beginning of each session, but also one week before (WB) the sessions, one day after (day5), and one week after (WA) the sessions

### Study design

The study is an interventional, randomized controlled trial. Participants were allocated randomly to one of two groups. The randomization script was generated in MATLAB by an independent third party (not involved in the study’s execution or analysis), who accordingly assigned participants to the corresponding group. The outcomes assessors were masked to the allocation of participants. In Group I (VR+tSTIM Group), participants experienced a combined visuo-tactile VR and targeted neurostimulation. Group II (VR Group) received only the VR intervention without any electrical stimulation. The required sample size was determined using G*Power, based on an effect size of 1.58 for TENS intervention compared to sham TENS in the treatment of neuropathic pain ^18^. An alpha error probability (α) of 0.05 and a statistical power of 0.8 were chosen. The calculation indicated a minimum of 8 participants per group. Except for the type of intervention provided, the experimental protocol was the same for both groups (Fig. 1D). One week before (WB) the beginning of the intervention sessions, participants completed pain questionnaires (Neuropathic Pain Symptom Inventory (NPSI) ^36^, and Visual Analogue Scale (VAS) ^37^ (Fig. 1D). Participants received the pain intervention for four consecutive days. NPSI and VAS were collected at the beginning of each session to assess the pain of the previous 24 hours. The intervention consisted of 20 minutes of VR+tSTIM or VR, depending on the experimental group. Before and after the pain intervention, an EEG baseline session of three minutes was recorded (Fig. 1D). To assess the impact of the intervention on sensory performances, the 2PD Test ^38^ and the Proprioceptive Displacement Test ^39^ were performed on the first and last day (Fig. 1D). As a follow-up, participants completed VAS and NPSI questionnaires one day (DA) and one week (WA) after the end of the intervention (Fig. 1D).

### tSTIM calibration

In the VR+tSTIM group, at the beginning of each session, tSTIM calibration was performed to find the optimal stimulation parameters for the four nerves of interest (tibial and peroneal nerves of both feet) using an interactive custom-made GUI. Participants were instructed to describe the intensity of the stimulation they felt on a 0-10 scale. 0 is not perceived, 1 is barely perceivable, and 10 is a very strong, not bearable sensation ^80,81^. The tSTIM was delivered with biphasic rectangular charge-balanced pulses, at a frequency *f* = 50 *Hz* as previously used in other studies ^82–84^. For each nerve, an initial stimulation ramp (biphasic pulses, of increasing amplitude (A) and fixed pulsewidth (*PW* = 300*μs*)) was delivered until the participants reported a sensation of 5/10 intensity. If the elicited sensation was perceived as somatotopic (spreading along the nerve and matching the desired foot location), then electrodes position and stimulation amplitude were saved, otherwise, electrodes were repositioned until the optimal location was achieved. As a second step, a calibration procedure similar to the one reported in ^39,80,81^ was conducted to find the perceptual (minimum, 2/10 intensity) and maximum threshold (below pain, 8/10 intensity) for each nerve for each participant. Participants repeated the ramps of fixed A and increasing PW three times. The mean of these PW values across the three repetitions was saved as minimum and maximum PW to modulate the electrical stimulation in the real-time intervention. On the following days, the position of the electrodes, A, and PW of the previous days were used as the starting point of the calibration. Then, when the sensations were not somatotopic or the intensity was not correct, the previous calibration steps were repeated to achieve an optimal, somatotopic sensation.

### Pain assessments

As a primary outcome, the Neuropathic Pain Symptom Inventory (NPSI) ^36^ was used. The NPSI is the standard validated tool to assess neuropathic pain severity ^85^. It is specifically validated to evaluate the different symptoms of neuropathic pain for a 24-hour interval. It is scored as the sum of 10 pain descriptor items (rated on a scale from 1 to 10 each). Together with NPSI, we collected the VAS of participants a complementary pain rating^37^. For VAS, participants indicated their pain on a straight line with one end meaning no pain and the other end meaning the worst pain imaginable.

### Sensory assessments

To measure proprioception, participants underwent the Proprioceptive Displacement Test (Fig. 3A) ^39^. During the test, they sat on a chair and placed their foot on a raised support. While the foot was covered (using a panel) participants were asked to verbally move the pole of a 3D-printed measurement device until it matched the position of their hallux. This was repeated *N* = 10 times by repetitively changing the initial position of the foot. The mean error between the reported and correct hallux position was defined as the outcome measure. In other words, the less is the displacement, the more correct is the body representation of the lower limb. To investigate the impact of the intervention of tactile acuity, the 2PD Test ^38^ was performed (Fig. 3B). While blindfolded, participants were repetitively touched (*N* = 20) with either one or two pins at a fixed distance in the plantar and dorsal sides of the foot and asked to determine the correct number of pins ^86,87^. The percentage of success was taken as the outcome measure. Pin distance was individually calibrated before the first session by gradually increasing the pin distance from 10 mm with 5 mm steps. For each distance, participants were touched five times with both pins and were asked to identify the number of pins (one or two). The distance at which participants correctly identified at least two out of five stimuli was selected. The sensory tests were repeated twice in the protocol, on the first and last day of the intervention

### EEG analysis

Before and after every pain intervention session, participants underwent three minutes of baseline EEG recording. For each recording, EEG signals were band-pass filtered (0.5 − 42 *Hz*) with a windowed-sinc Hamming Finite Impulse Response (FIR) filter of order 2000. Independent Component Analysis was performed to remove eye and muscular artifacts. Then, data was re-referenced to the average of all 24 channels. The analysis was conducted only on the central (Cz, C3, C4, CPz, CP1, CP2) and parietal (Pz, P1, P2) regions, given their key role in cortical pain processing^40^. For each channel, the Power Spectral Density (PSD) distribution was estimated with the Welch Method (*window size* = 2*s*, *overlap* = 50%). Mean PSD, absolute and relative spectral power were then computed for four bands of interest: Delta (1 − 4 *Hz*), Theta (4 − 8 *Hz*), Alpha (8 − 13 *Hz*), Beta (13 − 30 *Hz*), and Gamma (30 − 40 *Hz*). Once computed, features were standardized for each participant across all sessions. To evaluate objective correlates of the intervention, we examined changes in EEG features from the initial to the final day of the intervention, utilizing the baseline recording post-pain intervention on both occasions. Additionally, to investigate the associations between EEG features and reported pain, we conducted a correlation analysis that integrated variations in EEG features with participants’ changes in the NPSI score.

### Statistical analysis

In each analysis, we selected either parametric or non-parametric tests based on the normality of the data, as determined by the Shapiro tests. For the pain analysis, NPSI and VAS decrease among the experimental group was evaluated using clinically significant threshold ^41^, rather than with statistical tests. Clinical significance was defined as a 30% and 2/10 points (for VAS, 20/100 for NPSI) reduction, while substantial clinical significance was defined as 50% reduction i.e., the thresholds considered to define responders endpoints in clinical trials for pain treatment ^41,42,45^. Between groups, independent t-tests (or Mann-Whitney tests, depending on the normality) were used to compare the NPSI and VAS decrease at each timepoint. For the 2PD and Propioceptive displacement tests, the statistical analysis was carried among groups, using the paired t-test (or the Wilcoxon test, depending on the normality). Similarly, the EEG analysis of feature variation between the first and the last day of intervention was conducted among groups using the paired t-test (or the Wilcoxon test, depending on the normality). The correlation between EEG features variation and reported pain variation was carried out with Pearson’s correlation coefficient (or Spearman’s rank correlation coefficient, depending on the normality).

## RESULTS

Eighteen participants (10 females, 8 males, mean age of 66 ± 12) with chronic neuropathic pain participated in the study (See CONSORT diagram, Fig. S1) from July 2022 to July 2024. Of these, thirteen participants developed neuropathic pain as a complication of peripheral diabetic neuropathy; two participants had painful idiopathic neuropathy; two participants had Drug-Induced Peripheral Neuropathy (DIPN) and one participant was diagnosed with Tarsal Tunnel Syndrome (TTS) neuropathy (Table 1).

**Table 1.** Participants’ characteristics. DPN = Diabetic Peripheral Neuropathy; DIPN = Drug-Induced peripheral neuropathy, TTS = Tarsal tunnel syndrome. PC01-PC08 are intended as control patients.

|  | Age (years) | Height (cm) | Weight (kg) | Origin of Neuropathy | Time since diagnosis (years) | Experimental condition |
| --- | --- | --- | --- | --- | --- | --- |
| P01 | 76-80 | 175 | 75 | DPN | 15 years | VR+tSTIM |
| P02 | 61-65 | 180 | 82 | DPN | 11 years | VR+tSTIM |
| P03 | 76-80 | 151 | 59 | DPN | 9 years | VR+tSTIM |
| P04 | 66-70 | 189 | 96 | DPN | 4 years | VR+tSTIM |
| P05 | 76-80 | 180 | 80 | DPN | 16 years | VR+tSTIM |
| P06 | 46-50 | 180 | 68 | DIPN | 2 years | VR+tSTIM |
| P07 | 56-60 | 163 | 50 | DIPN | 2 years | VR+tSTIM |
| P08 | 56-60 | 174 | 61 | DPN | 25 years | VR+tSTIM |
| P09 | 76-80 | 182 | 85 | Idiopathic Neuropathy | 5 years | VR+tSTIM |
| P10 | 76-80 | 168 | 90 | DPN | 30 years | VR+tSTIM |
| PC01 | 56-60 | 190 | 111 | DPN | 13 years | VR |
| PC02 | 71-75 | 190 | 95 | DPN | 23 years | VR |
| PC03 | 56-60 | 170 | 68 | TTS | 2 years | VR |
| PC04 | 56-60 | 165 | 55 | Idiopathic Neuropathy | 1 year | VR |
| PC05 | 71-75 | 159 | 94 | DPN | 40 years | VR |
| PC06 | 51-55 | 178 | 95 | DPN | 26 years | VR |
| PC07 | 76-80 | 176 | 67 | DPN | 25 years | VR |
| PC08 | 46-50 | 158 | 70 | DPN | 8 years | VR |

The multisensory platform offered synergistic pain intervention temporally synchronizing VR and targeted neurostimulation (Fig. 1A). Participants wore a headset showing a relaxing beach scenario from a first-person perspective. The VR waves (approaching and retracting to participants’ feet) were coupled to modulated tSTIM targeting the tibial and peroneal nerves (Fig. 1B, Video S1). Stimulation parameters and locations were individually calibrated to optimize nerve activation, eliciting a somatotopic sensation that spreaded distally from the stimulation site across the entire foot. Participants were randomly allocated to the VR+tSTIM group (receiving coupled VR+tSTIM stimulation) or VR group (exposed to VR only) (Fig. 1C) and underwent four consecutive days of 20 minutes intervention (Fig. 1D). Statistical analyses between the two groups (VR and VR+tSTIM) were performed to ensure comparability regarding age, NPSI scores, and VAS scores (Table S2). To gather participants’ pain reports, NPSI, and VAS were daily collected starting from one week before the intervention to one week after it (Fig. 1D). Detailed characterization of participants’ pain profiles, along with specific parameters of the tSTIM and VR interventions, can be found in Table S3 and Fig. S2. The sensory effect of the multimodal platform was assessed on the first and last intervention day with the Two-Point Discrimination Test and Proprioceptive Displacement Test (Fig. 1D). EEG recordings were daily collected to measure objective neurophysiological indicators of treatment efficacy (Fig. 1D).

### VR+tSTIM intervention clinically reduces neuropathic pain

The participant’s subjective pain analysis was conducted following Dworkin et al. ^41^ recommendations, hence using two different methods to evaluate the clinical importance of pain improvement. For NPSI, clinically significant thresholds were therefore set at 30% and 20 points reductions from the baseline NPSI (average between one week before and D1 NPSI). Similarly, VAS thresholds were 30% and 2 points reduction respectively.

Participants in the VR+tSTIM group (*N* = 10) showed clinically significant (according to both thresholds) mean NPSI reduction persisting from the third day 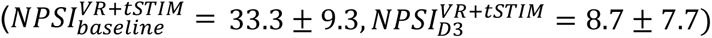 to one week after the ending of the intervention 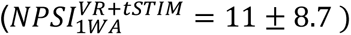(Fig. 2A)(Table S4). Single-participant analysis showed a substantial (higher than 50%^41–43^) NPSI decrease from baseline to one week after in 7/10 participants and a moderate pain decrease (more than 30%^41–43^) in 1/10 participant. In the VR group, (*N* = 8), the mean NPSI variations across days were not clinically different from the baseline NPSI (Fig. 2B)(Table S4). Only one participant showed substantial (>50%) NPSI decrease from baseline to one week after the intervention. We then conducted a between-group analysis using independent t-tests (or Mann-Whitney, depending on normality) to compare the absolute pain variation between groups. A significant reduction in NPSI was observed in the VR+tSTIM group compared to the VR group across all intervention days (Fig. 2C, Table S5).

**Figure 2.**
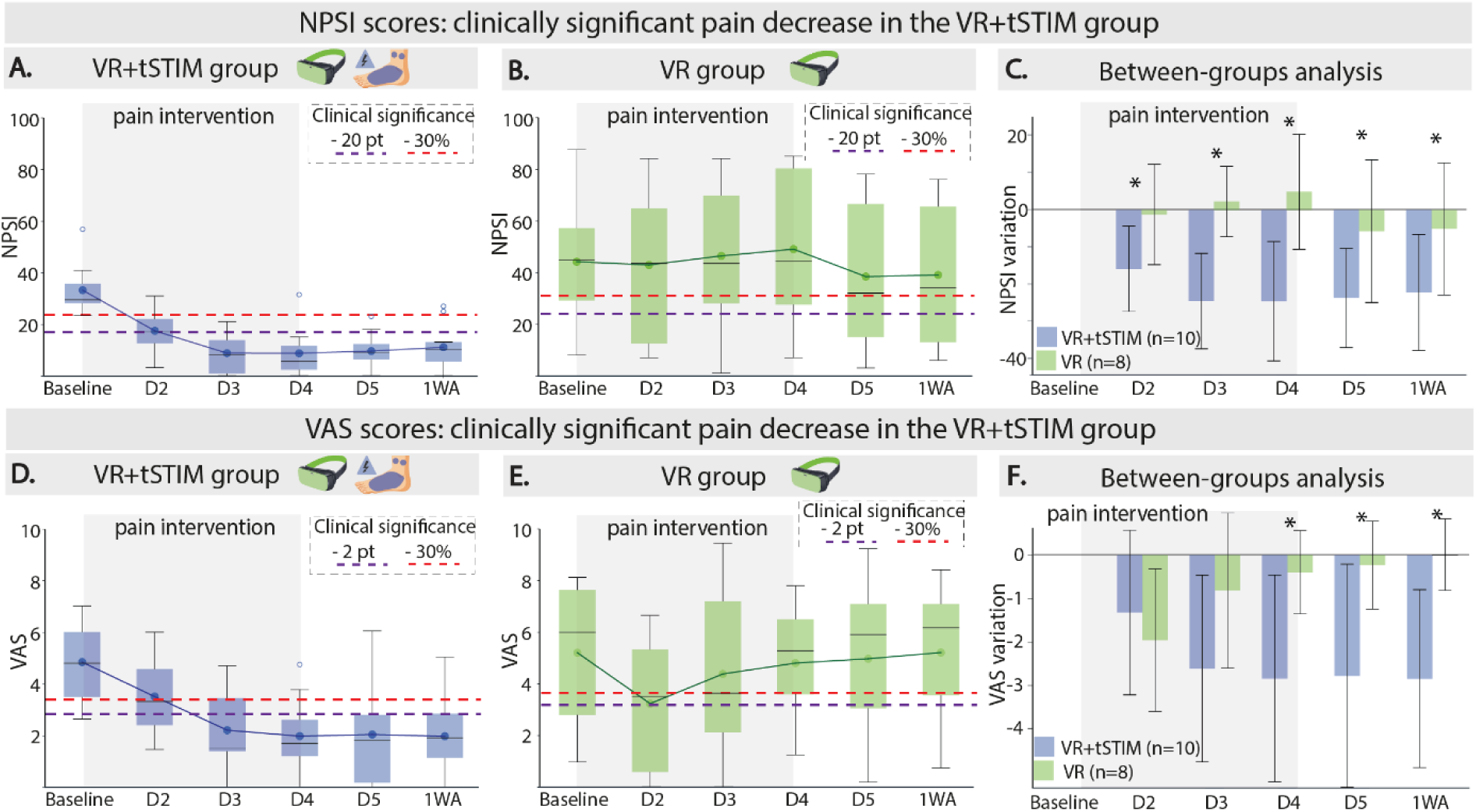
NPSI and VAS pain changes over the intervention. A-B) NPSI distribution for VR+tSTIM (A) group and VR group (B). Boxplots of NPSI are shown for each day. For each boxplot, the line and the dot correspond to the median and mean of the distribution, respectively. The red (−30%) and purple (−20pt) lines show clinically significant pain reduction. C) NPSI pain reduction from baseline. For each day, pain reduction of VR+tSTIM (blue), and VR group (green) are shown as barplots (mean ± std). T-test or Mann-Whitney tests (depending on normality) were used for statistical analysis. E-F) VAS boxplot distribution for VR+tSTIM (E) group and VR group (F). G) VAS pain reduction from baseline. For C) and G): * p < 0.05, ** p < 0.01, *** p < 0.001. Acronymous: D1, D2, D3, D4: Day 1, Day 2, Day 3, Day 4 of the treatment; D5, 1WA: One day and one week after the treatment end respectively.

Similar results were observed for the VAS analysis. Participants in the VR+tSTIM group showed clinically significant mean VAS reduction lasting from the third day of intervention 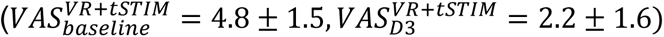 to the week after 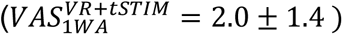(Fig. 2D)(Table S6). Single-participant analysis pointed at a substantial (higher than 50%) VAS decrease from baseline to one week after in 6/10 participants and a moderate pain decrease (more than 30%) in 2/10 participants. In the VR group, reported VAS was diminished on the second day 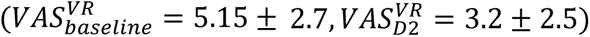, but only according to one of the criteria used (<30%)(Fig. 2E). No significant variations were observed in the upcoming days and at the follow-up (Fig. 2E) (Table S6). Similar to NPSI, only one participant showed moderate pain decrease (higher than 30%) at the one-week follow-up. The between-group analysis of absolute VAS variation indicated a significantly greater pain reduction in the VR+tSTIM group compared to the VR group from Day 4 through the one-week follow-up (Fig. 2F, Table S7).

### VR+tSTIM intervention improves tactile acuity and proprioceptive performance

Propioceptive displacement and tactile acuity, defined through the accuracy during two-point discrimination (2PD) test, were measured at the first and last therapy session, for both the VR+tSTIM and the VR group. Proprioceptive displacement, measured as mean error during 10 repetitions, is shown in Fig. 3C. The VR+tSTIM group showed a statistically significant reduction of the proprioceptive error (from 3.89±1.75 to 2.50±1.61, p=0.006). VR group showed no improvement (2.40±1.01 to 3.15±1.78 p=0.151). Tactile acuity, measured as correct trials in the 2PD tests, is shown in Fig. 3D. Tactile acuity statistically significantly improved from pre to post therapy for the VR+tSTIM group (55.4±19.2 to 66.1±13.7, p=0.012) but not for the VR group (45.6±28.1 to 47.6±26.6, p=0.423).

**Figure 3.**
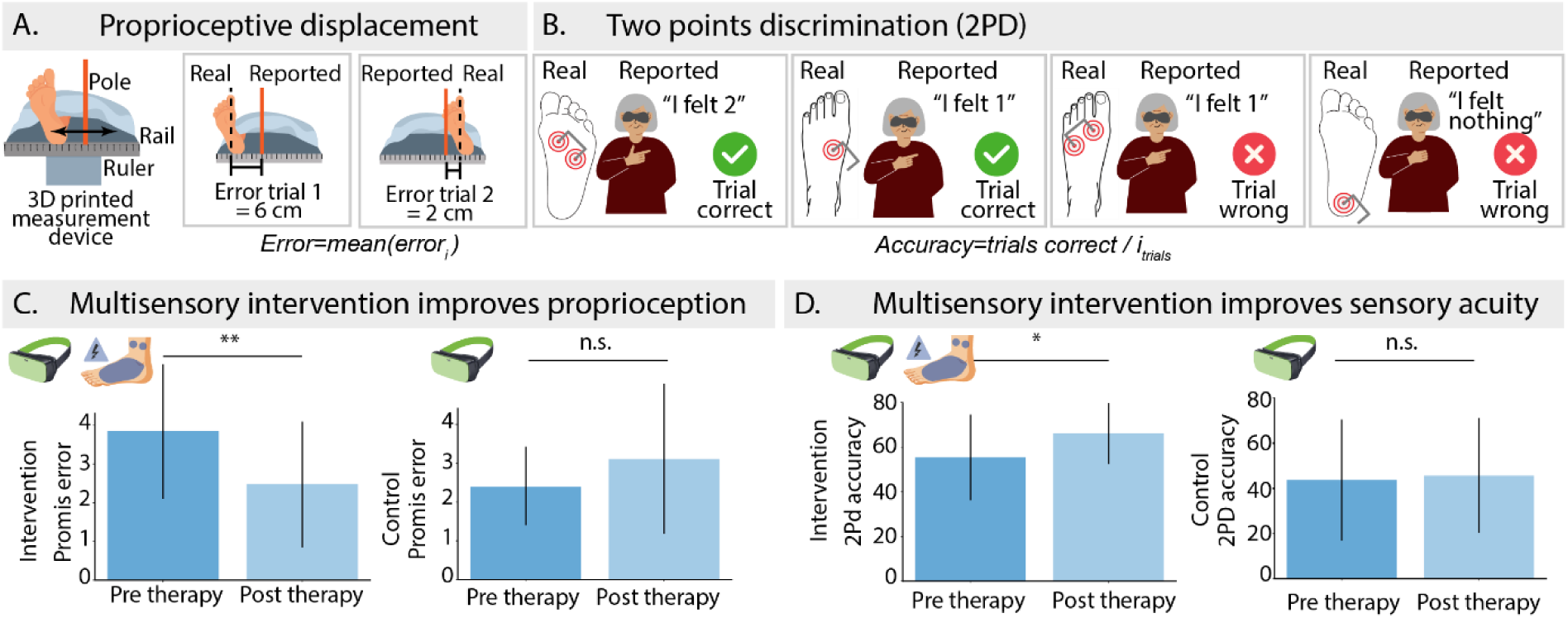
Sensory tests. A) Proprioceptive displacement protocol. The participant is sitting down with the leg covered. The displacement is defined as the spatial difference between the reported position and the real position of the hallux. B) Two points discrimination protocol. While blindfolded, participants are repetitively touched (N = 20) with either one or two pins at a fixed distance and report the perceived number of pins. The percentage of success is taken as the outcome measure. C) Proprioceptive displacement error for VR+tSTIM and VR group on the first and last day of intervention. D) Two points discrimination accuracy for the VR+tSTIM and VR group on the first and last day of intervention. Mannwhitney U test, * p<0.05, ** p<0.01. n.s.: not significant.

### Objective EEG digital biomarkers provide evidence for pain reduction

To find objective biomarkers supporting the therapeutic efficacy of the intervention, we observed participants’ neurophysiological EEG changes. Specifically, we computed the difference in EEG features between the first and the last day of intervention. We extracted band-power features representing the EEG information in the frequency domain. The EEG power analysis in the central and parietal areas highlighted the delta (δ), alpha (α) and gamma (γ) bands as the EEG indicators most affected by the pain intervention (Table S8). Three representative features of the overall behaviour are shown in Fig. 4. EEG relative delta power in the central region significantly decreased only after the VR+tSTIM intervention 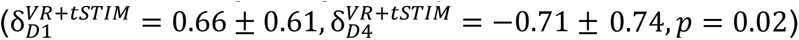 (Fig. 4A). This does not occur in the VR group, where the same indicator significantly increases 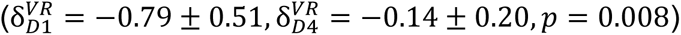 (Fig. 4A). Moreover, the change in feature value (from the first to the last day of the intervention) positively correlates with participants’ NPSI score changes (both in VR+tSTIM and VR group) (*Spearman ρ* = 0.66, *p* = 0.004) (Fig. 4A, Table S9). By performing separate linear regression analysis between the two groups, the slope of the intercept line was higher in the VR+tSTIM (*slope*_*VR*+*tSTIM*_ = 0.20) group compared to the VR group (*slope*_*VR*_ = 0.12) (Fig. 4A). The VR+tSTIM intervention also resulted in significant increase in EEG α PSD in central region 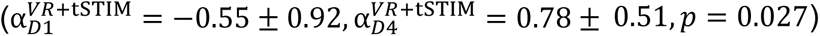 (Fig. 4B). Again, this was not seen in the VR group 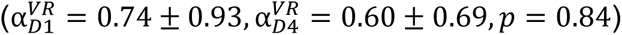 (Fig. 4B). The correlation analysis showed a significant negative correlation between the features and participants’ NPSI score changes (both in VR+tSTIM and VR group) (*Spearman ρ* = −0.65, *p* = 0.005) (Fig. 4B, Table S9). Separated linear regression showed lower slope for the VR+tSTIM (*slope*_*VR*+tSTIM_ = −0.61) intercept compared to the VR intercept (*slope*_*VR*_ = −0.08) (Fig. 4B). In the γ band, a significant decrease of EEG absolute parietal γ power was observed in the VR+tSTIM group only 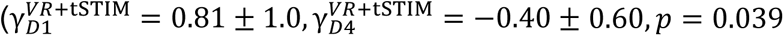; 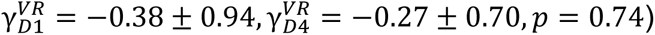. The feature showed a significant positive correlation with NPSI score changes (*Pearson ρ* = 0.65, *p* = 0.005) (Fig. 4C, Table S9), and again a higher slope for the VR+tSTIM group (*slope*_*VR*+tSTIM_ = 1.20) compared to the VR group (*slope*_*VR*_ = 0.30) in the separate linear regression analysis.

**Figure 4.**
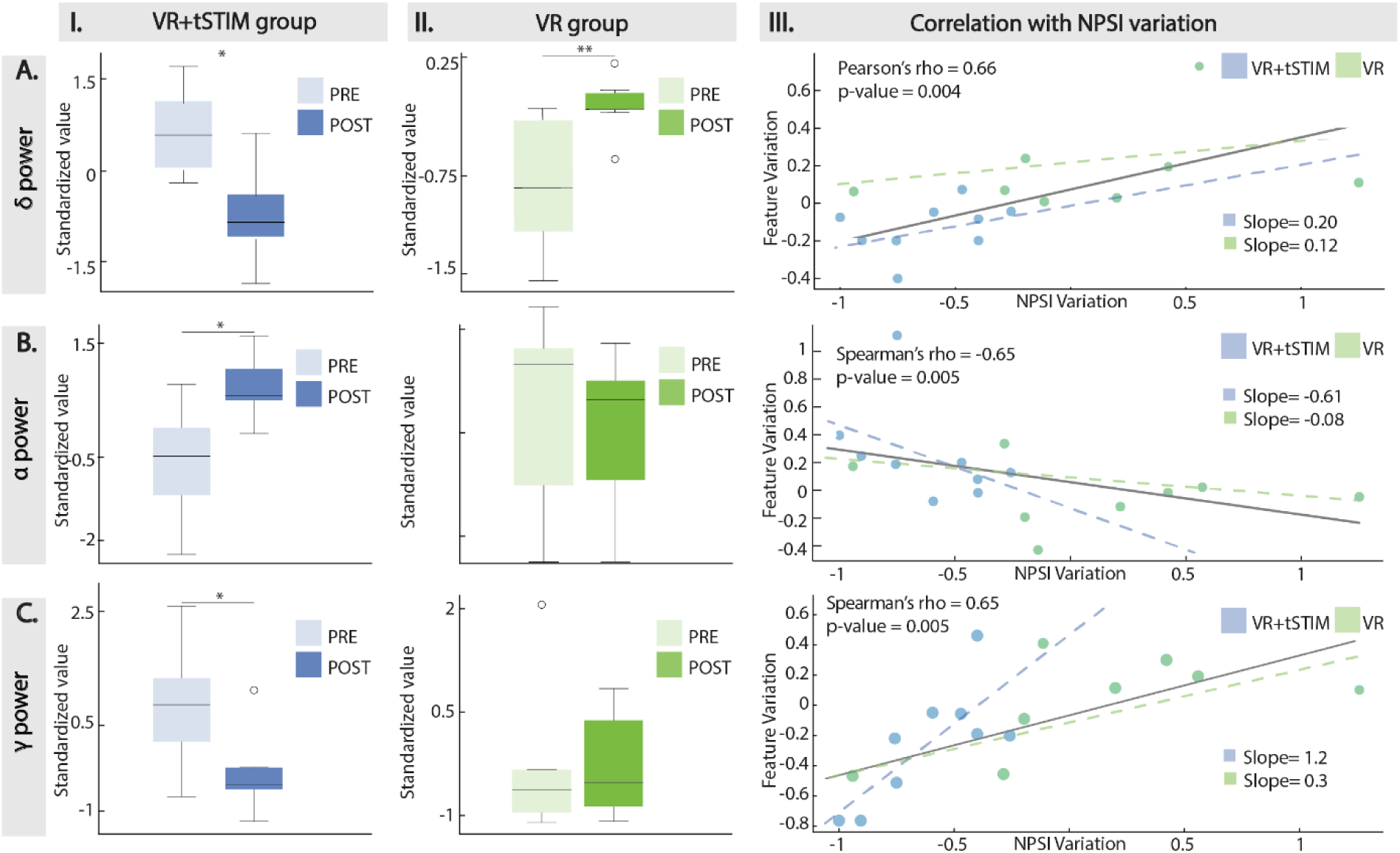
Neurophysiological indicators of pain. A) EEG delta power. Variation of EEG relative delta power in the central region from D1 to D4 for the VR+tSTIM group (I) (N = 9) and the VR group (II) (N = 8). In III, the correlation between the feature variation and the NPSI variation for all participants is shown (grey line). The correlation coefficient and p-value are reported. The correlation of the VR+tSTIM (in blue) and VR (in green) group are also shown. The slopes of the respective linear regressions are reported. B) EEG alpha power. Variation of EEG PSD in the central region from D1 to D4 for the VR+tSTIM group (I) and the VR group (II) and correlation analysis (III). C) EEG delta power. Variation of EEG gamma absolute power in the parietal region from D1 to D4 for the VR+tSTIM group (I) and the VR group (II) and correlation analysis (III). For I. and II.:Wilcoxon or paired t-test were used depending on normality. * p < 0.05, ** p < 0.01.

## DISCUSSION

Pain is complex and its multidimensional nature requires targeted treatments. The overuse and crisis of generic analgesic opioids highlights the need for alternative non-pharmacological solutions that address the multidimensionality of pain. However, it has been challenging to demonstrate therapeutic effectiveness of pain therapies due to the lack of reliable objective indicators of therapeutic response ^32,44^. To these aims, we developed a non-invasive, targeted VR-neurostimulation platform and evaluated its analgesic efficacy in an RCT. The targeted VR-neurostimulation was controlled with an active comparator comprising VR intervention only. We measured the intervention efficacy on comprehensive outcomes, including self-reported pain, sensory measures, and objective neural correlates of pain. Our results show that participants in the VR+tSTIM group experienced clinically significant reductions in self-reported pain, which were supported by improvements in sensitivity, proprioceptive tests, and therapy-induced changes in specific pain-related EEG power bands.

### Self-reported pain analysis

First, we showed that targeted VR neurostimulation clinically decreases pain as measured both through the NPSI and VAS scores. Clinical significance is defined as a 30% and 2/10 points (for VAS, 20/100 for NPSI) reduction in pain, which are the thresholds considered to define responders endpoints in clinical trials for pain treatment ^41,42,45^. The clinically significant reduction was achieved on the second (NPSI) and third day (VAS) of treatment, showing a cumulative effect and persisting up to the follow-up one-week post-intervention. In contrast, the VR intervention did not lead to any significant or lasting reduction in NPSI across the treatment days. However, participants in the VR group did report a significant reduction in VAS scores on the second day of intervention, a change that was not reflected in the NPSI results. This temporary improvement in VAS scores is likely due to the differences in how pain is measured by these two scales. VAS is a simple one-dimensional scale and is more susceptible to attentional factors and momentary biases ^46^, such as initial optimism about the therapy. In contrast, NPSI is a more comprehensive and robust assessment of neuropathic pain, spanning across different neuropathic pain subscales. Therefore, this transient VAS reduction may suggest a short-term attentional modulation effect of VR on pain ^25,47^, though lacking persistence.

After demonstrating the within-subject pain reduction of the targeted VR neurostimulation intervention, it was crucial to evaluate its efficacy against the VR group. The comparison with an active comparator is often missing in previous trials ^31,48,49^ and is essential for validating the true therapeutic benefit of the proposed intervention ^50^. Studies that only report within-group changes fail to establish whether a treatment is effective, as observed changes might result from factors other than the treatment itself (e.g., contextual biases) ^32,50^. We demonstrated that the targeted VR neurostimulation significantly decreases self-reported NPSI and VAS pain scores across treatment days and at follow-up, compared to VR-only. This finding showed that the multimodal intervention consistently outperformed the VR-only comparator, ruling out possible placebo effect of the application of a generic intervention.

Another key shortcoming of existing TENS or VR interventions is the lack of continuous monitoring of pain (typically assessed only post-intervention). This fails to provide information about pain evolution throughout the days of the treatment and the optimal duration needed for effective results ^18,25^. Our study design, by daily collecting multiple data, minimized recall bias and maximized ecological validity ^51^, while gaining valuable insights into patients’ pain dynamics and determine the minimum duration of the intervention for significant. Our findings indicate that two days of VR+tSTIM intervention are required to reach clinically significant improvements, as verified by two different pain measurements ^41^. Additionally, the analgesic effects of four consecutive days of intervention persist for at least one-week post-treatment. However, to precisely determine the optimal dosing and duration of therapy, further research with a larger sample size and a longer follow-up is needed.

### Targeted multisensory intervention

While our study validated the benefits of multisensory interventions for neuropathic pain, it remains unclear whether the observed pain reduction was due solely to the optimized targeted neurostimulation or its combined effect with VR. However, we can partially attribute the strong analgesic effects to our neurostimulation approach, which improves upon conventional TENS methods for treating lower-limb neuropathic pain.

Indeed, most clinical studies employ standardized, non-personalized TENS parameters, delivering pre-defined waveforms to all participants without accounting for individual variations in neuropathy severity and sensitivity. For instance, ^48^ and ^52^ used fixed parameters (amplitude and pulse width of the stimulation) for all participants regardless of patients’ perception. Furthermore, the literature generally adopts a non-personalized approach to electrode placement, typically describing it as “close to the pain site” ^18^. For example, ^53^ used four electrodes on the vastus medialis, vastus lateralis, fibula, and gastrocnemius; ^52^ positioned electrodes near the dorsum pedis and at the top of the fibula; and ^48^ applied two electrodes on the upper shin and above the ankle. Moreover, none of these studies adjusted electrode placement based on individual differences. In contrast, we performed a targeted personalized calibration process to determine both the intensity of stimulation and the optimal electrode positioning. Amplitude and pulse width were adjusted to provide clearly perceivable electrical stimulation for each participant, later modulated to match the VR visual input. Electrode placement was optimally calibrated to achieve a somatotopic sensation distally spreading on the foot. This, differently from electro cutaneous stimulation providing localized sensations, ensured the activation of the large diameter sensory fibers in the nerves responsible for neuropathic pain ^21^, amplifying the natural effects of the gate control theory in pain inhibition. The advantages of targeted stimulation are further supported by fMRI studies, showing that targeted somatotopic neurostimulation successfully generates somatosensory sensations in the cortex of neuropathic participant ^54^. In contrast, in-loco electro cutaneous stimulation at the pain site often fails to activate the sensory cortex ^54^. This suggests that, due to the neuropathic damage, such stimulation may not effectively engage the peripheral large-diameter fibers and the ascending sensory pathways crucial for spinal cord inhibition as described by the gate control theory ^19^.

### Tactile and proprioceptive readouts of pain

Alongside self-reported pain measures, objective sensory pain readouts were collected. Diminished tactile acuity has been reported in peripheral neuropathies^55^, due to lesions of the somatosensory pathways disrupting the transmission of tactile stimuli, and in different chronic pain conditions ^56^. Interestingly, tactile acuity, assessed using the 2PD test, showed significant improvement in the VR+tSTIM group following the intervention. A combination of peripheral and central factors can explain this finding ^56,57^. Several studies demonstrated that TENS plays a role in increasing blood circulation ^58–61^, underscoring its potential impact on nerve damage ^62^. Therefore, the provided electrical stimulation (received by the VR+tSTIM group only) may directly influence the integrity of sensory pathways and contribute to improved acuity. At the central level, extensive research showed changes in representational fields in the primary somatosensory cortex associated with alterations in 2PD thresholds^63–67^. Our findings may reflect neuroplastic adaptations in the representational areas of the somatosensory cortex following the VR+tSTIM intervention. These neuroplastic changes likely enhance the brain’s ability to process tactile information more accurately, thus concurrently improving tactile acuity and reducing pain perception. This central perspective could also explain the improvement observed in the proprioceptive displacement test, where participants in the VR+tSTIM group demonstrated an enhanced ability to accurately locate the position of their lower limb. The proposed task tests for participants’ short-term body representation, (i.e. current limb angles and positions) as encoded in their sensorimotor cortex ^27^. Evidence indicates that neuropathic pain is associated with structural changes in the somatosensory cortex ^63,68^, and generates anomalous body perception (e.g. body parts perceived as “heavy”, “constricted” or “swollen”)^39,69–71^. In this context, multisensory stimulations (especially visuo-tactile contingencies) have been advocated as effective therapeutic tools for altering the subject’s sense of ‘self’ ^26^ and directly impacting body representation to alleviate pain ^28–30,72^. Hence, the observed results are an indicator of the impact of the VR+tSTIM intervention on network dynamics^73^ to effectively modulating body representation. The lack of improvement in the VR condition can be explained by the necessity for congruent stimuli from multiple modalities to create a robust and coherent bodily illusion ^74^. Since the VR group received only visual stimulation, the same effect on restoring and improving body representation was not observed.

### Neurophysiological indicators of pain

To assess pain changes, self-reported and sensory pain readouts need to be coupled electrophysiological pain indicators, i.e., biomarkers, to objectively monitor therapeutic response and disease progression ^32,35^. EEG is a non-invasive and cost-effective method for gathering neurophysiological pain data. While the search for EEG chronic pain biomarkers is extensive ^40,75^, most studies are either cross-sectional, comparing chronic pain patients to healthy subjects, or descriptive studies monitoring patients undergoing a single intervention or no intervention ^75^. Our study design not only enables the monitoring of EEG correlates of chronic pain, but also allows for the independent assessment of neural responses to two different interventions, serving as objective indicators of the efficacy of each intervention. We observed a significant reduction in EEG gamma and delta and an increase in alpha power bands, pre- to post-treatment, exclusively in the VR+tSTIM group. Gamma activity is crucial in pain processing, with several chronic pain studies reporting increased gamma as a potential marker of pain ^75^. Similarly, the alpha power is a key biomarker in the modulation of chronic pain, with higher alpha power associated to chronic pain conditions ^40,75^. Delta waves are typically associated with acute pain conditions, with increased delta activity indicating a painful stimulus or condition ^76^. While the VR+tSTIM intervention provided an analgesic effect accompanied by reduced delta power, delta power increased following the VR intervention only. This aligns with ^77^, who observed a short-term delta power increase after a pain VR intervention and suggested it reflects a cognitive modulation effect of the immersive VR experience.

Additionally, we correlated these objective responses with participants’ self-reported pain. In simpler words, we analyzed whether the reduction in pain as perceived subjectively by the participants was supported by objective neural changes. Delta, gamma, and alpha bands changes correlated with NPSI pain reduction. This suggests that the greater the reduction in pain, the higher the difference in the neural biomarker from pre- to post-therapy. When no reduction in pain was observed (as often for the VR group), the relative neural biomarkers were not showing significant changes. Notably, the slope of the fitted linear regression was steeper (in absolute value) for the VR+tSTIM group compared to the VR group. This indicates a stronger modulatory effect of the multisensory intervention in altering these key pain-related EEG bands compared to the VR intervention. Overall, our results provide clear evidence that the multisensory intervention effectively modulates neural pathways of pain, supporting the efficacy of the targeted VR-neurostimulation treatment.

### Limitations and future perspective

While this is the first RCT on a multimodal VR-neurostimulation pain intervention, the current study also presents some limitations. While we can support that VR+tSTIM has a strong effect on pain both subjectively and objectively, and that VR alone does not affect it, we cannot conclude whether the observed results are attributable solely to the neurostimulation or its combination with VR visual input. The VR condition primarily served to mitigate the influence of participants’ predispositions and momentary biases toward treatment success, ensuring that the results (especially the subjective ones) were not merely due to optimistic expectations and placebo effects. Future studies should aim to compare the multisensory intervention against tSTIM-only stimulation. To rigorously test the hypothesis of brain plasticity and changes in body representation following the multisensory intervention, future studies should incorporate high-resolution brain imaging techniques (e.g., fMRI). These methods can evaluate connectivity and structural indexes of the somatosensory cortex following pain interventions. Future studies should also aim to incorporate additional wearable devices, such as wristbands, to further identify pain-related neurophysiological correlates. This approach will advance the development of real-time, portable digital technologies for monitoring therapeutic responses through objective indicators of pain patterns.

## Acknowledgments

The authors are immensely grateful to the volunteers who freely donated their time to the advancement of knowledge and to a better future for pain treatments. The authors thank Jothini Sritharan, Stefan Fuchs, Jonas Schröder for their help in experiments, and Dr. med. Bogdan Mimor for helping with recruitment. The funder had no role in the experimental design, analysis, or manuscript preparation or submission. All authors had complete access to data. All authors authorized submission of the manuscript, but the final submission decision was made by the corresponding author.

## Author contributions

G.V.A. designed the platform, performed all the experiment and all the analyses, made the figures and wrote the manuscript; N.G. performed all the experiment and all the analyses, made the figures and wrote the manuscript; M.W. and N.B. contributed to experiments and data analysis; G.P. helped in the experiments and designed the study; N.S. helped in the experiments; C.Z. recruited participants, discussed the protocol and results; S.R. designed the study, supervised the experiments, supervised the analyses, discussed the results and wrote the manuscript. All the authors authorized submission of the manuscript, while the final submission decision was taken by the corresponding author.

## Data and materials availability

All completely anonymized data, code, and materials used in the analysis are available upon reasonable request from the corresponding author.

## Supplementary materials and results

**Figure S1.**
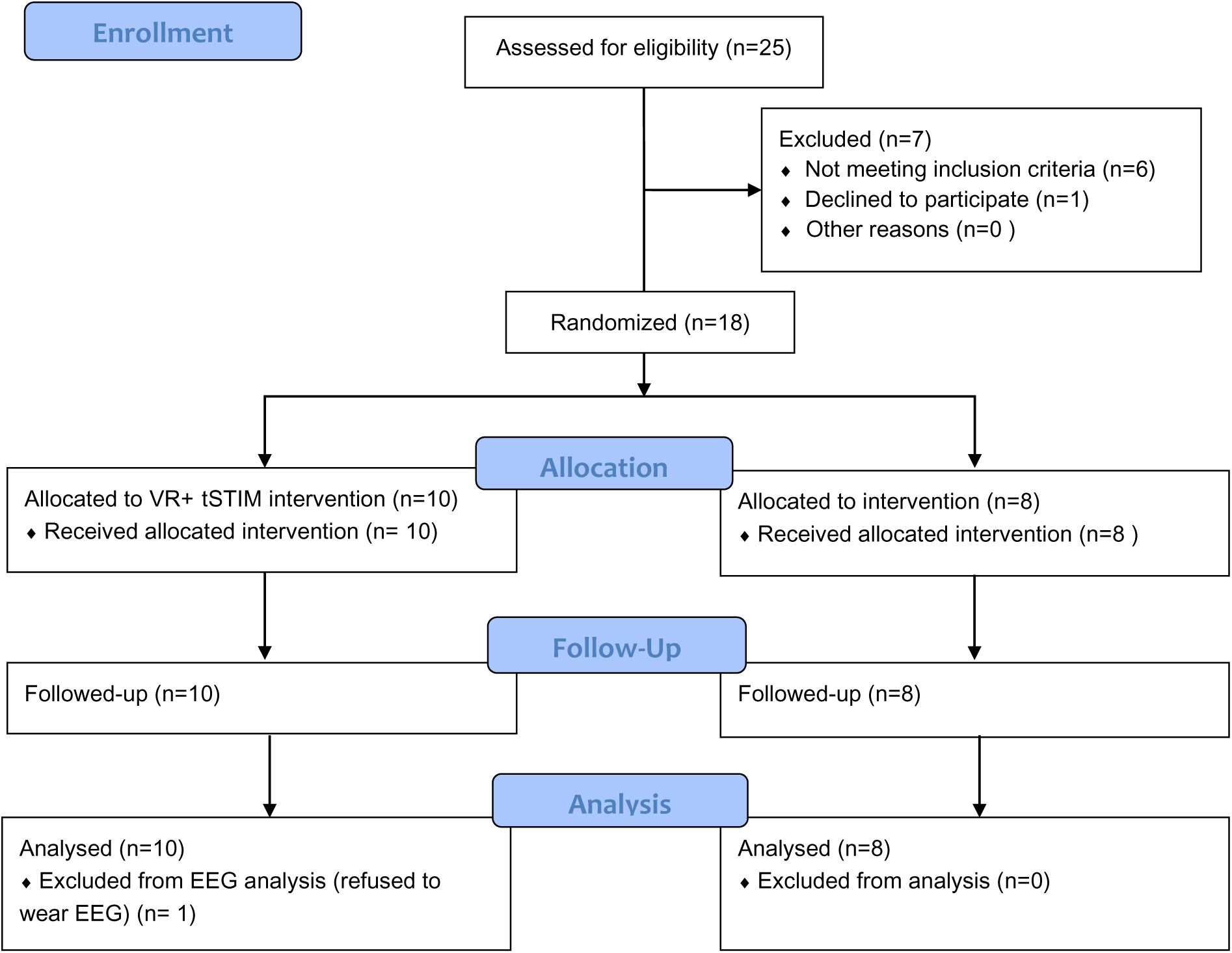
CONSORT diagram for RCT.

**Figure S2.**
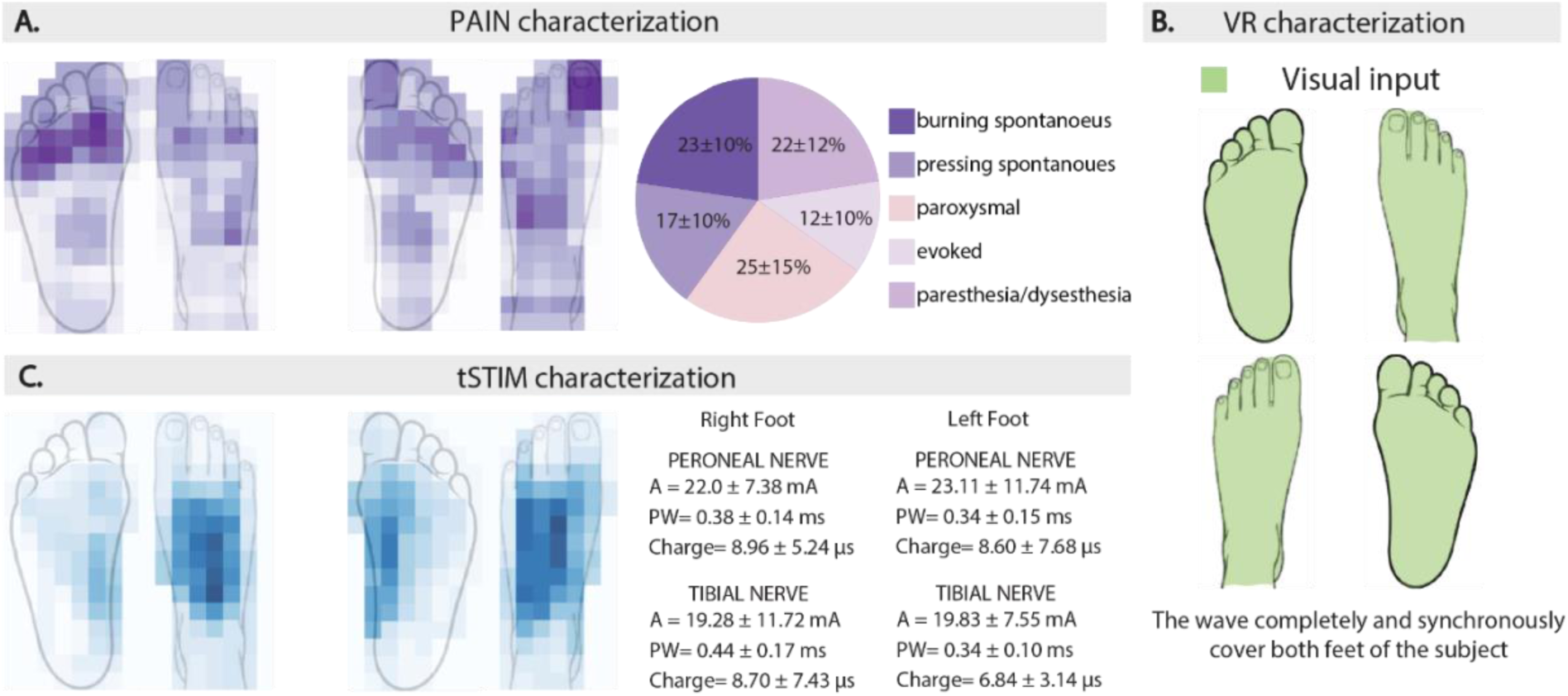
Pain, tSTIM, and VR characterization. A) Pain Characterization. On the left, the location where the patients felt pain (the more intense the color, the higher the number of patients who reported pain in that area). On the right, a pie-chart of the subscales of patients’ baseline NPSI. B) VR characterization. The visual wave covered both feet of the avatar. B) tSTIM Characterization. On the left, the location where the patients felt the electrical stimulation. On the right, tSTIM values (*mean* ± *std* across patients).

**Table S1.**
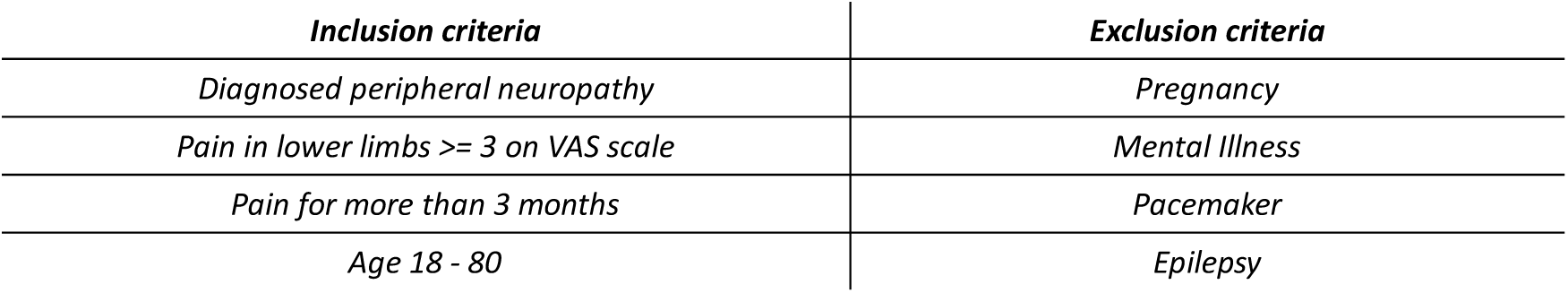
Inclusion and exclusion criteria.

| *Inclusion criteria* | *Exclusion criteria* |
| --- | --- |
| *Diagnosed peripheral neuropathy* | *Pregnancy* |
| *Pain in lower limbs $\geq 3$ on VAS scale* | *Mental Illness* |
| *Pain for more than 3 months* | *Pacemaker* |
| *Age 18 - 80* | *Epilepsy* |

**Table S2.** Grouped baseline analysis. Age, BMI, and baseline NPSI and VAS statistical analysis across groups.

|  | *VR+tSTIM*<br>*(mean ± std)* | *VR*<br>*(mean ± std)* | *pvalue* |
| --- | --- | --- | --- |
| *Age* | 68 ± 11.8 | 62.3 ± 10.9 | 0.306 |
| *BMI* | 24.5 ± 3.8 | 27.2 ± 5.5 | 0.233 |
| *NPSI* | 33.3 ± 9.8 | 44.5 ± 24.9 | 0.305 |
| *VAS* | 4.8 ± 1.57 | 5.2 ± 2.9 | 0.750 |

**Table S3.** tSTIM parameters. For each VR+tSTIM patient, tSTIM Amplitude (A), maximum Pulsewidth (PW), and Charge (C) are reported for dorsal (D) and plantar (P) stimulation of both feet (RF, LF). Each patient’s A, PW, and C values correspond to the mean across the four days of stimulation.

|  | *A*<br>*D_RF*<br>*(mA)* | *PW*<br>*D_RF*<br>*(μs)* | *C*<br>*D_RF*<br>*(nC)* | *A*<br>*P_RF*<br>*(mA)* | *PW*<br>*P_RF*<br>*(μs)* | *C*<br>*P_RF*<br>*(nC)* | *A*<br>*D_LF*<br>*(mA)* | *PW*<br>*D_LF*<br>*(μs)* | *C*<br>*D_LF*<br>*(nC)* | *A*<br>*P_LF*<br>*(mA)* | *PW*<br>*P_LF*<br>*(μs)* | *C*<br>*P_LF*<br>*(nC)* |
| --- | --- | --- | --- | --- | --- | --- | --- | --- | --- | --- | --- | --- |
| *P01* | 30 | 550 | 16.5 | 45 | 600 | 27 | 46 | 490 | 22.5 | 21 | 550 | 11.6 |
| *P02* | 25 | 590 | 14.8 | 22 | 700 | 15.4 | 27 | 600 | 16.2 | 22 | 450 | 9.9 |
| *P03* | - | - | - | 8 | 600 | 4.8 | - | - | - | 11 | 425 | 4.7 |
| *P04* | - | - | - | 8 | 600 | 4.8 | - | - | - | 25 | 340 | 8.5 |
| *P05* | 15 | 295 | 4.4 | 15 | 322.5 | 4.8 | 15 | 332.5 | 5 | 20 | 277.5 | 5.6 |
| *P06* | 25.8 | 367.5 | 10.4 | 23 | 365 | 8.9 | 21.5 | 282.5 | 6.1 | 31.5 | 330 | 10.1 |
| *P07* | 8.8 | 275 | 2.4 | 7.3 | 287.5 | 2.1 | 9 | 290 | 2.6 | 7.3 | 232.5 | 1.6 |
| *P08* | 24.5 | 335 | 8.2 | 25.8 | 335 | 8.7 | 24.5 | 265 | 6.5 | 25 | 290 | 7.3 |
| *P09* | 25 | 242.5 | 6.1 | 26.3 | 250 | 6.6 | 18.8 | 182.5 | 3.5 | 23.8 | 215 | 5.2 |
| *P10* | - | - | - | 12.5 | 310 | 3.9 | - | - | - | 11.8 | 337.5 | 4.1 |
| *ALL* | 22 ± 7.4 | 379.3 ± 136.8 | 9 ± 5.3 | 19.3 ± 11.7 | 437 ± 167 | 8.7 ± 7.4 | 23.1 ± 11.7 | 348.9 ± 144.8 | 8.9 ± 7.5 | 19.8 ± 7.5 | 344.8 ± 103.8 | 6.9 ± 3.2 |

**Table S4.** Patients’ NPSI values. Acronymous: D1, D2, D3, D4: Day 1, Day 2, Day 3, Day 4 of the treatment; 1WB, D5 5, 1WA: One week before, one day and one week after the treatment end respectively.

|  | *1WB* | *D1* | *D2* | *D3* | *D4* | *D5* | *1WA* |
| --- | --- | --- | --- | --- | --- | --- | --- |
| *P01* | 35 | 32 | 26 | 20 | 15 | 13 | 13 |
| *P02* | 30 | 30 | 10 | 8 | 10 | 18 | 12 |
| *P03* | 41 | 41 | 23 | 13 | 2 | 10 | 8 |
| *P04* | 30 | 17 | 11 | 0 | 4 | 9 | 0 |
| *P05* | 34 | 39 | 19 | 14 | 2 | 0 | 0 |
| *P06* | 28 | 28 | 17 | 3 | 7 | 7 | 13 |
| *P07* | 27 | 31 | 31 | 21 | 31.5 | 23 | 25 |
| *P08* | 51 | 63 | 17 | 0 | 0 | 6 | 5 |
| *P09* | 35 | 15 | 17 | 0 | 12 | 9 | 27 |
| *P10* | 29 | 29 | 3 | 8 | 3 | 0 | 7 |
| *PC01* | 28 | 49 | 32 | 34 | 37 | 35 | 35 |
| *PC02* | 51 | 51 | 55 | 53 | 52 | 3 | 6 |
| *PC03* | 27 | 8 | 13 | 19 | 31 | 18 | 14 |
| *PC04* | 83 | 92 | 82 | 84 | 79 | 74 | 76 |
| *PC05* | 74 | 50 | 84 | 84 | 84 | 78 | 76 |
| *PC06* | 66 | 45 | 59 | 65 | 85 | 64 | 62 |
| *PC07* | 11 | 5 | 11 | 1 | 7 | 6 | 10 |
| *PC08* | 33 | 33 | 7 | 31 | 17 | 29 | 33 |

**Table S5.** NPSI variations and p-values. Baseline is the average between week before and Day 1. Acronymous: D1, D2, D3, D4: Day 1, Day 2, Day 3, Day 4 of the treatment; 1DA, 1WA: One day and one week after the treatment end respectively.

| | $\Delta NPSI_{VR+tSTIM}(\text{mean} \pm \text{std})$ | $\Delta NPSI_{VR}(\text{mean} \pm \text{std})$ | *p-values* |
| --- | --- | --- | --- |
| *Baseline* | - | - | - |
| *D2* | $-15.85 \pm 10.93$ | $-1.25 \pm 12.66$ | 0.025 |
| *D3* | $-24.55 \pm 12.19$ | $2.25 \pm 8.4$ | <0.001 |
| *D4* | $-24.6 \pm 15.24$ | $4.88 \pm 14.49$ | 0.001 |
| *1DA* | $-23.75 \pm 12.66$ | $-5.75 \pm 17.97$ | 0.032 |
| *1WA* | $-22.25 \pm 14.83$ | $-5.13 \pm 16.64$ | 0.026 |

**Table S6.** Patients’ VAS values. Acronymous: D1, D2, D3, D4: Day 1, Day 2, Day 3, Day 4 of the treatment; 1WB, D5, 1WA: One week before, one day and one week after the treatment end respectively.

|  | 1WB | D1 | D2 | D3 | D4 | D5 | 1WA |
| --- | --- | --- | --- | --- | --- | --- | --- |
| P01 | 4.585 | 4.57 | 5.095 | 3.43 | 2.81 | 1.4745 | 2.3055 |
| P02 | 4.95 | 4.95 | 4.905 | 4.335 | 4.715 | 4.95 | 5 |
| P03 | 5.815 | 8.1 | 5.955 | 4.665 | 0.665 | 0.15 | 3 |
| P04 | 6.975 | 5.05 | 3.17 | 0 | 1.19 | 6 | 3 |
| P05 | 2.615 | 4.03 | 1.46 | 0.32 | 0 | 0.255 | 0.165 |
| P06 | 3.89 | 3.89 | 3.43 | 1.43 | 1.875 | 2.895 | 1.28 |
| P07 | 3.015 | 2.23 | 3.17 | 3.4 | 3.75 | 2.435 | 2.24 |
| P08 | 5.9649 | 5.63 | 2.1 | 1.375 | 1.93 | 2.135 | 1.0975 |
| P09 | 3.125 | 3 | 3.43 | 1.5 | 1.5 | 0 | 1.555 |
| P10 | 7.5 | 6.25 | 2.125 | 1.5 | 1.25 | 0 | 0 |
| PC01 | 5.33 | 3.615 | 4.9 | 2.48 | 4.5 | 4.915 | 5.565 |
| PC02 | 1.345 | 2.79 | 0 | 2.833 | 1.5 | 0.185 | 1.39835 |
| PC03 | 1.15 | 0.7625 | 0.7625 | 0.9325 | 1.229 | 0.282 | 0.726 |
| PC04 | 9.8335 | 5 | 3.1935 | 4.37 | 5.9665 | 7 | 6.6665 |
| PC05 | 8.051 | 8.051 | 6.592 | 6.9145 | 6.423 | 9.153 | 8.354 |
| PC06 | 7.5225 | 8.075 | 6.4175 | 7.82 | 7.735 | 6.805 | 6.875 |
| PC07 | 3 | 3 | 0 | 0 | 4.24 | 3.94 | 4.24 |
| PC08 | 7.5 | 7.5 | 3.75 | 9.375 | 6.5 | 7.125 | 7.5 |

**Table S7.** VAS variations and p-values. Baseline is the average between week before and Day 1. Acronymous: D1, D2, D3, D4: Day 1, Day 2, Day 3, Day 4 of the treatment; D5, 1WA: One day and one week after the treatment end respectively.

| | $\Delta VAS_{VR+tSTIM}(\text{mean} \pm \text{std})$ | $\Delta VAS_{VR}(\text{mean} \pm \text{std})$ | p-values |
| --- | --- | --- | --- |
| Baseline | - | - | - |
| D2 | $-1.32 \pm 1.79$ | $-1.96 \pm 1.53$ | 0.465 |
| D3 | $-2.61 \pm 2.04$ | $-0.82 \pm 1.67$ | 0.076 |
| D4 | $-2.84 \pm 2.26$ | $-0.40 \pm 0.90$ | 0.015 |
| D5 | $-2.78 \pm 2.44$ | $-0.23 \pm 0.95$ | 0.018 |
| 1WA | $-2.84 \pm 1.94$ | $0.01 \pm 0.77$ | 0.002 |

**Table S8.**
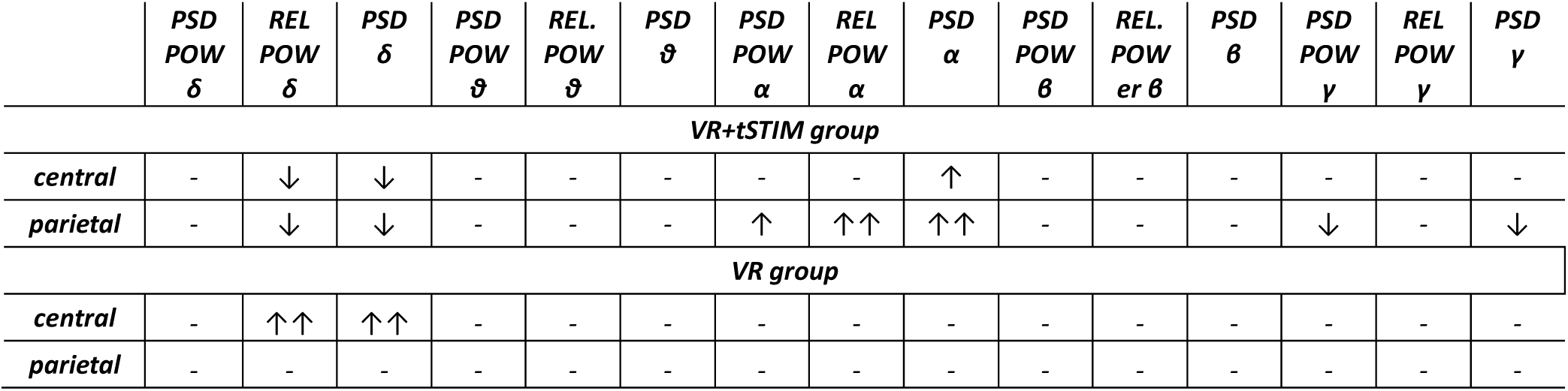
EEG features statistical analysis. Upward arrows indicate that the distribution of that specific feature significantly increases from day 1 to day 4. Downward arrows indicate the opposite. Depending on normality, independent t-test or Mann-Whitney test were used (↑↑: p <= 1.00e-02; ↑: p <= 0.05; -= non-significant).

**Table S9.**
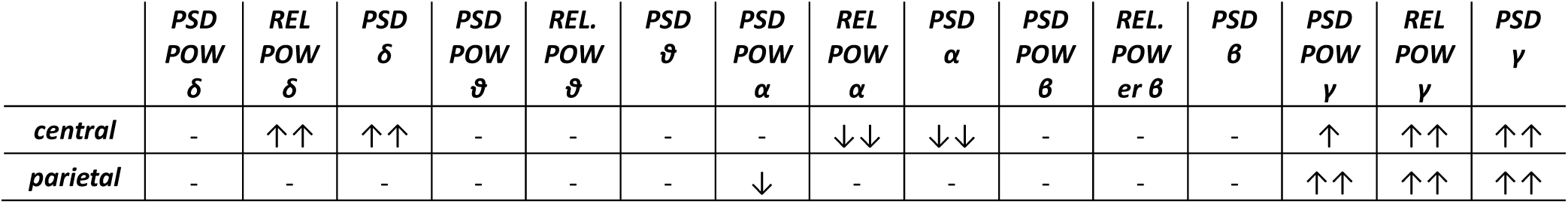
EEG features - NPSI correlation analysis. Upward arrows indicate a positive correlation between feature value and NPSI variation from day 1 to day 4. Downward arrows indicate the opposite. Depending on normality, Pearson or Spearman correlation were used (↑↑: p <= 1.00e-02; ↑: p <= 0.05; -= non-significant).

